# Effects of Parietal Cathodal tDCS during Game Cue Exposure on Internet Gaming Disorder: A Randomized Double-Blind Sham-Controlled Trial

**DOI:** 10.1101/2025.07.20.25331859

**Authors:** Yixuan Song, Yuchen Huang, Qihong Zheng, Xiaoqin Yang, Yang Guo, Huixing Gou, Junjie Bu, Tianye Jia, Guangdong Zhou, Jie Shi, Yan Sun

## Abstract

**Background:** Internet Gaming Disorder (IGD) is officially listed as a behavioral addiction, exhibits high prevalence and has inadequate treatment efficacy. Targeting craving triggered by gaming cues represents a critical therapeutic objective. This study aimed to establish optimizing neuro-electrophysiologic biomarkers for IGD and develop a targeted neuromodulation protocol.

**Methods:** In an exploratory study, we identified the optimized electroencephalography (EEG) indicators of IGD diagnose and craving through machine learning models based on event-related potential (ERP) during game cue exposure across two independent datasets (D1: 25 IGD, 22 Recreational Game Users (RGU), and 28 non-gaming Healthy Controls (HC); D2: 23 IGD and 23 HC). In an intervention study, we conducted a randomized, double-blind trial in 46 IGD participants, comparing active versus sham transcranial direct current stimulation (tDCS) targeting the optimized EEG marker. Active stimulation (1.5 mA, 20 min, 2 days) was applied during cue exposure (cathode: Pz; anode: right trapezius), while sham mimicked initial/final ramping without sustained current. The primary outcome was game craving (measured by QGU-B, VAS, and craving during exposure to presented/unpresented gaming cues) and daily gaming time, measured post-intervention and at 1 to 4 weeks follow-ups. The trial was registered at ClinicalTrials.gov (NCT06759051).

**Findings:** Parieto-occipital P300 (with maximal discriminative power at central parietal (Pz), IGD>HC) during game reactivity emerged as optimized EEG indicators for IGD discrimination (accuracy > 80%), and were associated with craving. Then, Pz targeted cathodal tDCS synchronized with game cue exposure could significantly reduce craving (p < 0.001), gaming time (p < 0.001), and P300 alpha (p=0.048) after intervention and at 1 to 4 weeks follow-ups, with concomitant improvement of decision-making in the active group. Crucially, treatment effects could be generalized to novel gaming cues.

**Conclusions:** These findings advance precision biomarkers and evidence-based neuromodulation strategies for IGD.

## Introduction

As the global video game market is expected to hit $522 billion in 2025 and esports becomes an official sport in events like the Asian Games, the growing accessibility and social acceptance of gaming have heightened public health concerns about problematic gaming use.^1^ Internet Gaming Disorder (IGD) is defined by persistent and uncontrolled gaming behaviors that result in clinically significant impairment across physical, psychological, and social functioning domains.^2^ Diagnostically, while the DSM-5 classifies IGD as a condition requiring further research, the ICD-11 formally codifies gaming disorder as an addictive disorder.^3,4^ A hallmark pathological feature of IGD is maladaptive cue-induced craving, driving persistent symptoms and relapse.^5^ Affected individuals also demonstrate cognitive deficits, such as impaired impulse control, blunted sensitivity to real-world rewards, and dysfunctional decision-making.^6,7^ Epidemiological data reveal a global IGD prevalence of 6.7% (2024 meta-analysis), with males showing approximately 2.5-fold higher rate than females.^8^ Despite its high prevalence and frequent psychiatric comorbidities, IGD frequently goes undiagnosed and untreated. However, evidence-based treatments specifically targeting IGD remain underdeveloped.^9^

Neuroscience-informed neuromodulation has emerged as a promising approach for IGD treatment. Cue-induced craving, a core IGD feature, can be objectively measured through electroencephalography (EEG). For instance, game cues could elicit distinct event-related potentials (ERP) patterns directly linked to prefrontal dysfunction and impaired cognitive control.^10^ Also, EEG studies have revealed mixed neural oscillatory patterns in IGD under cue exposure in the parieto-occipital regions.^11^ Furthermore, occipital alpha/beta oscillations during cue exposure may correlate with symptom severity in IGD.^12^ These EEG signatures enable machine learning (ML) applications, which could effectively distinguish IGD from healthy controls by integrating EEG and clinical features, offering promising biomarkers for craving and severity assessment of IGD.^13^ Therefore, advances in EEG monitoring and neuromodulation technologies now enable identification of IGD-specific neuro-electrophysiological signatures, which could not only elucidate the neural mechanisms underlying IGD, but also establish an evidence base for developing precisely targeted neuromodulation interventions.

Transcranial direct current stimulation (tDCS) alters cortical excitability via polarity-dependent membrane potentials changes: anodal stimulation increases excitability by depolarization, whereas cathodal decreases it by hyperpolarization. This technique shows promise for treating addictive behaviors.^14^ However, while dorsolateral prefrontal cortex (DLPFC)-targeted neuromodulation has proven effective in enhancing executive control and reducing substance consumption and craving in substance use disorders (e.g., alcohol, nicotine, and drugs), its application to IGD remains limited with inconsistent craving reduction outcomes.^15–18^ These mixed findings thus highlight the necessity to identify IGD-specific neuro-electrophysiological biomarkers to optimize tDCS protocols.

Therefore, this study pursued two primary objectives: firstly, to identify EEG biomarkers for IGD diagnosis and craving using ML, and secondly, to evaluate intervention effects of tDCS targeting these identified neural signatures. We acquired resting-state and cue-elicited EEG data from individuals with IGD, recreational game users (RGU, which represents a “risk group” between addicted and healthy states), and non-gaming healthy controls (HC), alongside assessments of gaming severity and craving. The ML model was trained and validated across two independent datasets to detect the optimized cortical EEG features linked to IGD diagnosis and craving. Subsequently, we conducted a randomized, double-blind, sham-controlled trial for subjects with diagnosed IGD to assess whether tDCS modulation of the neural activity of proposed could reduce craving and ameliorate cognitive dysfunction. Together, these findings establish a framework for biomarker-guided diagnosis and precision neuromodulation in IGD.

## Methods

### Study design

There were two parts in this study. In the exploratory study (part 1), we identified optimized electroencephalography (EEG) indicators of IGD diagnosis and craving through an EEG-based ML model during game cue exposure across two independent datasets. In the intervention study (part 2), the therapeutic potential of tDCS on IGD targeting the optimized EEG indicator (derived from the ML model) was assessed by conducting a randomized, double-blind, controlled trial.

This study was reviewed and approved by the Peking University Institutional Review Board (IRB00001052-24146) and was performed following the relevant guidelines and regulations in two cities in China (Beijing and Tianjin). The randomized, double-blind, controlled trial was also registered with ClinicalTrials.gov, NCT06759051.

### Participants

Subjects were recruited via online advertisements, including individuals with Internet gaming disorder (IGD), recreational game users (RGU), and healthy controls (HC). All subjects were required to be aged between 18 and 25 and signed an informed consent form and voluntarily participated in this trial.

Inclusion criteria of the IGD group comprise: 1) Familiar with and mainly play Honor of Kings; 2) Meet at least 6 entries in the DSM-5 diagnostic criteria for Internet gaming disorder and 3) Played online games for at least 14 h per week for the past 12 months so far.

Inclusion criteria of the RGU group comprise:1) Familiar with and mainly play Honor of Kings; 2) Meets the DSM-5 diagnostic criteria for Internet gaming disorder with the number of entries less than 5; and 3) Played online games for at least 14 h per week for the past 12 months so far.

Inclusion criteria of the HC group comprise:1) Familiar with Honor of Kings; 2) Meet the number of entries less than 5 in the DSM-5 diagnostic criteria for Internet gaming disorder; and 3) Played online games less than 7 h per week for the past 12 months so far.

Exclusion criteria comprise:1) Subjects who were diagnosed as mental disorders of schizophrenia spectrum disorder, bipolar disorder, depression, anxiety disorder, obsessive-compulsive disorder and neurocognitive dysfunction; 2) A history of addictive substance dependence; 3) History of comorbid serious organic diseases (including diabetes, cardiovascular diseases, abnormal thyroid function, malignant tumors and other clinically confirmed chronic diseases); 4) Use of psychoactive drugs (antidepressants, mood stabilizers, antipsychotics, etc.) within two weeks prior the enrollment; 5) Current use of drugs affecting the central nervous system (including antipyretic and analgesic drugs, tranquilizers, antibiotics and herbal preparations, etc.); 6) Medical conditions with persistent somatic symptoms (e.g., chronic pain syndromes, intractable itchy skin, etc.); 7) History of tDCS intolerance or skin sensitization; and 8) Inadequate study cooperation (3 or more missed appointments or less than 80% operational compliance during the assessment period).

The exploratory study consisted of two datasets: Discovery Dataset 1 (D1), all subjects were recruited from Tianjin between April 11, 2024 and June 20, including 25 in the IGD group, 22 in the RGU group, and 28 in the HC group,2024; Validation Dataset 2 (D2), the IGD group was all from Beijing, with a total of 23 subjects, and the HC group was all from Tianjin, with a total of 23 subjects. The subjects in intervention study were all IGD, and a total of 46 subjects were recruited from universities in Tianjin (n=23) and Beijing (n=23, the same participants in IGD group of D2), which began on December 30, 2024 and continued through February 15, 2025. (Figure 1)

**Figure 1:**
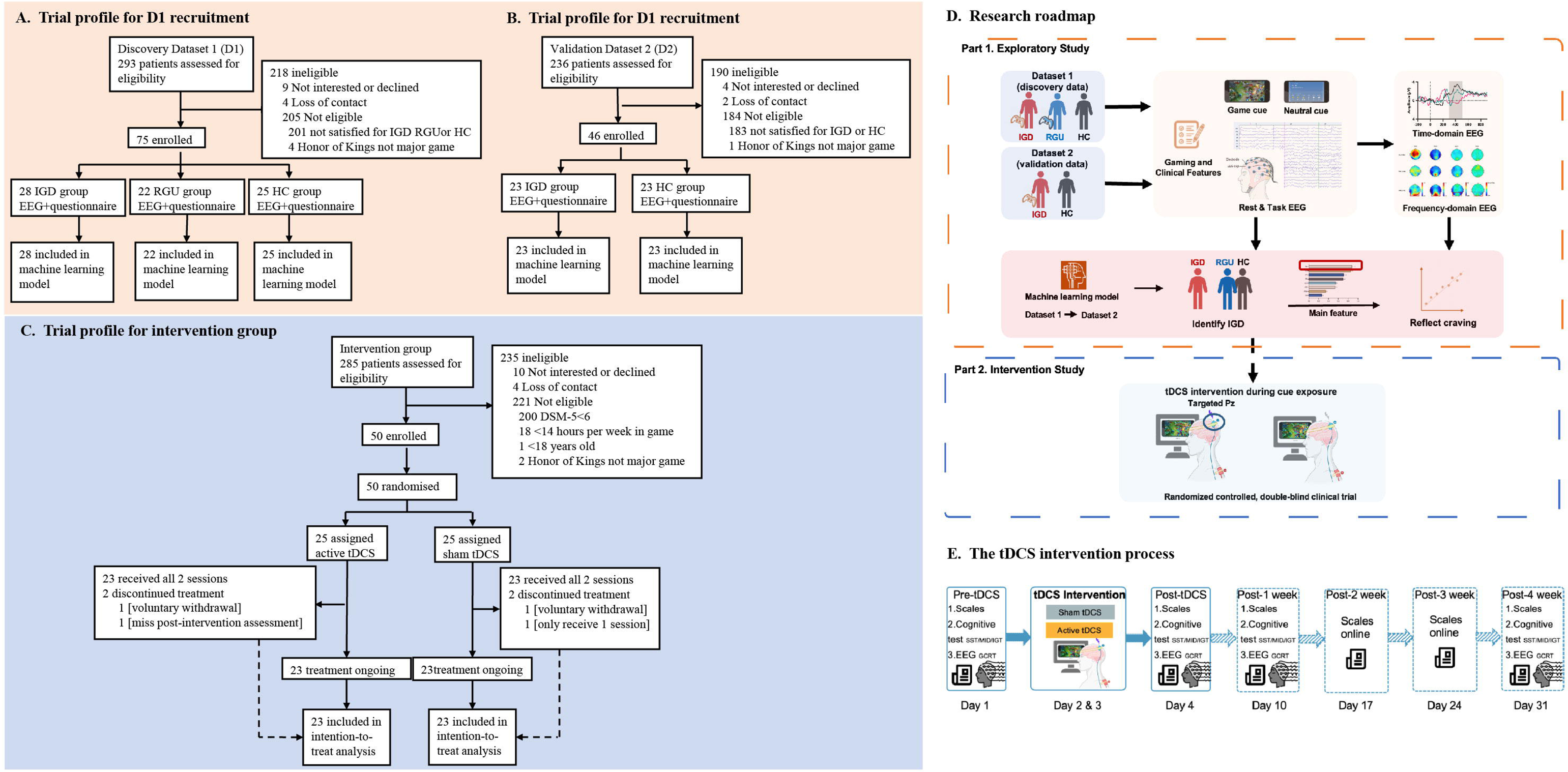
Trail profile and Experimental procedure. The exploratory study in D1 (A) and D2 (B). C: The intervention study. D: Research roadmap. E: The tDCS intervention process. IGD: Internet gaming disorder; RGU: recreational game user; HC: healthy control; tDCS: transcranial direct current stimulation.

### Randomisation and masking

Participants were randomly allocated (1:1) to active or sham tDCS groups using a computer-generated randomization sequence in Excel: each subject received a unique random number between 0 and 1 and was ranked accordingly, and group assignment (active=1, sham=0) was determined via modulo operation. This allocation was performed solely by one experimenter who generated subject-numbered stimulation files. Double-blinding was implemented: operators saw only subject-numbered files (no stimulation parameters) and selected files blindly. Sham stimulation replicated initial/terminal skin sensation without cortical effects. Blinding success was assessed post-intervention by comparing subject-reported current intensity perceptions between groups.

### Procedures

#### Demographic information and questionnaires

Demographic information collected included the subject’s age, gender, education, health status, medication use, and acceptance of weak current stimuli. Addiction severity degree was assessed by DSM-5 diagnostic criteria for IGD and the 9-item Internet gaming disorder scale-short-form (IGDS9-SF).^19^ Game craving was assessed by the Questionnaire of Gaming Urges-Brief (QGU-B) and the visual analogue scale (VAS) with a straight line with “0” at one end indicating “no craving at all” and “9” at the other end indicating “craving very much”.^20^

The depression symptoms and anxiety symptoms were assessed by the Patient Health Questionnaire-9 (PHQ-9) and the Generalized Anxiety Disorder-7 (GAD-7), respectively. The subject’s emotional state (positive and negative mood) was assessed using the Positive and Negative Affect Schedule (PANAS).^21^

#### Cognition paradigm testing

The following tasks were under EEG in this study:

1) The game cue reactivity task (GCRT) primarily tests the subjects’ craving level for game-related cues; The exploration part includes 30 positive game cue pictures, 30 negative game cue pictures, and 30 neutral pictures; the intervention part includes 50 positive game cue pictures and 30 neutral pictures. During the tDCS intervention, only positive game cue pictures are presented, with a total of 30 shown (presented cues). Notably, there were 20 positive game pictures which were unpresented in GCRT were used to assess the generation effects of the intervention on craving. 2) The stop signal task (SST) is a classical experimental paradigm that assesses attentional and inhibitory control by measuring an individual’s ability to inhibit an initiated action during a task; 3) The Monetary Incentive Delay (MID) task is a common paradigm for examining reward and punishment processing during anticipation and response; 4) The Iowa Gambling Task (IGT) is a test where people make choices from card decks with hidden risks and rewards. It measures how well someone makes real-life decisions involving uncertainty and consequences.^22–25^ For the more details of task paradigm, please see appendix pp 2.

#### Procedures of the exploratory study

To explore the optimized location and indicator for neuromodulation of IGD, the baseline data for three groups (IGD, RGU and HC) were collected, including demographic/gaming questionnaires, closed-eye resting-state EEG, and task-state EEG (only GCRT). EEG was acquired using a 32-channel mbt Smarting system (10-20 placement, impedance <5kΩ, 500Hz sampling) with participants being instructed to minimized movement/artifacts (Figure 1D).

#### Procedures of the intervention study

The intervention trial spanned four consecutive in-lab days plus follow-ups at weeks 1-4 (Figure 1E). Day 1 collected baseline data: demographic/gaming questionnaires, followed by closed-eye resting-state EEG and task-state EEG (including GCRT, SST, MID, and IGT). Days 2-3 delivered active/sham tDCS synchronized with game cues for 2 consecutive days; pre/post stimulation assessed game craving, mood, side effects, and sensation intensity. Day 4 repeated post-intervention questionnaires, resting EEG, and task EEG. Follow-ups occurred at weeks 1 and 4 (in-lab: craving/mood questionnaires, resting/task EEG) and weeks 2-3 (online: weekly gaming characteristics/craving/mood assessments) (Figure 1D, E).

tDCS was delivered using the DC-STIMULATOR MC (Neuroconn, Germany). The cathode (5×5 cm) was placed at Pz (parietal cortex), and a matching anode on the right trapezius muscle. Active stimulation consisted of a 20-minute protocol: 10-second ramp-up to 1.5 mA, 19 min 40 s at 1.5 mA, and 10-second ramp-down. Sham stimulation replicated the initial/final 20-second ramp-up/down sensations (to 1.5 mA and back to 0 mA), but maintained 0 mA current for the intervening 19 min 20 s, mimicking skin sensation without cortical modulation.^26^

### Outcomes

In the intervention study, the primary outcomes were craving reduction (measured by QGU-B, VAS, and craving during exposure to presented/unpresented gaming cues) and daily gaming time. Secondary outcomes included changes in P300 component alterations at the Pz electrode and cognitive task performance (SST, MID, IGT metrics). Adverse events were assessed during/ post tDCS, including fever, burns, development of drowsiness, and so on. Data collected in both Beijing and Tianjin were centrally assessed.

### Statistical analysis

#### Behavioral data analysis

Statistical analyses used GraphPad Prism 9 (significance: p<0.05, two-sided). Demographic variables (gender, age, playtime, QGU-B, VAS, DSM-5, IGDS9-SF, PHQ-9, GAD-7) were compared with ANOVA (three-group comparisons in D1) or t-tests (D2, intervention groups) for continuous data, and chi-square for categorical data. Age was included as a continuous covariate to control for potential confounding effects. Exploratory analyses employed two-way ANOVA for craving: Group (IGD/RGU/HC) × Cue-Type (Positive/Neutral/Negative Game Cues) with Šídák post-hoc. Pre-intervention analyses included: one-way ANOVA for cue-exposure craving (Neutral/Active-Presented/Active-Not-Presented cues) with Šídák post-hoc; independent t-tests for baseline SST (Go accuracy, Go RT, Stop accuracy, and SSRT) and MID metrics (RT/d’ across conditions) between active/sham groups. Longitudinal intervention effects used two-way RM-ANOVA with Group (active/sham) × Time (pre/post/1wk/4wk) for: mood (PANAS, PHQ-9, GAD-7), craving (QGU-B, VAS), playtime, SST/MID metrics, and IGT net scores across stages (6×10 trials), with Šídák post-hoc identifying significant timepoints and group differences.

#### EEG analysis

EEG Data were preprocessed in EEGlab/FieldTrip (MATLAB 2021b), excluding subjects with >20% rejected segments or PCA components. Resting-state EEG spectral analysis was performed using fast Fourier transform to extract signal frequency characteristics. The resulting power spectral density (PSD) was logarithmically transformed to yield decibel power spectral density (dBPSD). Time-frequency analysis employed the multitaper convolution method combined with a Hamming window, with results output as PSD. Frequency analysis (delta:1-4Hz, theta:4-8Hz, alpha:8-13Hz, beta:13-30Hz) and ERP analysis (P300:300-500ms) were performed. Group comparisons used repeated-measures ANOVA: for resting-state/P300, between-group factors were subgroups (D1: IGD/RGU/HC; D2: IGD/HC) with electrode sites as the within-group factor; for intervention effects on cue-exposure P300 at Pz, the between-group factor was treatment (active/sham) with time (pre/post/1-week/4-week) as the within-group factor (Šídák post-hoc tests). Spearman correlations examined associations between: 1) Pz P300 amplitude changes and craving changes post-intervention, and 2) baseline Pz P300-band power and post-intervention craving reduction.

#### Machine learning

For machine learning algorithms (part 1), Support Vector Machines (SVM) with a linear kernel (MATLAB 2021b) were used to classify IGD vs. HC/RGU groups by finding an optimal hyperplane. The model incorporated features from resting-state EEG (energy of four frequency bands across 31 electrodes) and the P300 time window (power spectral density at 8 electrodes), with a regularization parameter C optimized. Model performance was evaluated using: 1) leave-one-out cross-validation within datasets D1 or D2, and 2) training on D1 and testing on D2. Key evaluation metrics included accuracy, sensitivity (IGD identification), specificity (control identification), and AUC. Feature importance was analyzed by extracting SVM weight vectors across cross-validation rounds to identify high-contribution electrodes, followed by Spearman correlation analysis with craving metrics.

## Results

### Participant characteristics of the exploratory study

Age differed significantly among IGD, RGU and HC groups in D1 (F_(2,72)_ = 6.24, p = 0.003) and between IGD and HC groups in D2 (t_(44)_ = 3.14, p = 0.003). Sex distribution was comparable. All gaming measures, including daily gaming duration, QGU-B craving scores, IGDS9-SF scores, and DSM-5 criteria, showed significant group differences (all p < 0.001 in either D1 or D2). Besides, PHQ-9 and GAD-7 scores also differed significantly among groups in two datasets (all p<0.017) (Table 1).

**Table 1:** Demographic information and game use characteristics.

|  | Dataset 1 |  |  |  |  | Dataset 2 |  |  |  | Intervention groups |  |  |  |
| --- | --- | --- | --- | --- | --- | --- | --- | --- | --- | --- | --- | --- | --- |
|  | HC (n=28) | RGU (n=22) | IGD (n=25) | *p* | Statistics | HC (n=23) | IGD (n=23) | *p* | Statistics | Active (n=23) | Sham (n=23) | *p* | Statistics |
| Age (years) | 20.43 (0.50) | 20.59 (0.85) | 21.60 (2.00) | 0.0030 | F=6.24 | 20.43(0.10) | 22.04(0.50) | 0.0030 | t=3.14 | 20.78(0.41) | 21.09(0.51) | 0.643 | t=0.46 |
| Male/Female (Female%) | 12/16 (57.14%) | 11/11 (50.00%) | 13/12 (48.00%) | 0.78 | $\chi^2=0.49$ | 11/12(52.17%) | 12/11(47.83%) | 0.77 | $\chi^2=0.09$ | 9/14(39.13%) | 10/13(43.48%) | 0.768 | $\chi^2=0.09$ |
| Daily gaming time | 0.88 (0.22) | 3.59 (1.22) | 4.32(1.65) | <0.001 | F=64.33 | 0.89(0.04) | 2.90(0.21) | <0.001 | t=9.49 | 3.02(0.23) | 3.18(0.22) | 0.622 | t=0.32 |
| QGU-B | 26.75 (9.68) | 25.05 (8.99) | 46.52 (13.40) | <0.001 | F=29.73 | 26.44(1.89) | 47.30(1.63) | <0.001 | t=8.38 | 45.96(1.49) | 46.96(1.81) | 0.680 | t=0.42 |
| IGDS9-SF | 17.82 (4.18) | 18.64 (4.79) | 35.44 (3.07) | <0.001 | F=151.81 | 17.30(0.84) | 28.48(0.75) | <0.001 | t=9.90 | 31.52(1.31) | 29.13(1.18) | 0.152 | t=1.76 |
| VAS | NA | NA | NA | NA | NA | NA | NA | NA | NA | 6.09(0.28) | 6.35(0.27) | 0.507 | t=0.67 |
| DSM-5 | 1.86 (1.27) | 2.64 (2.01) | 7.16 (1.49) | <0.001 | F=82.64 | 1.70(0.24) | 6.78(0.20) | <0.001 | t=16.38 | 7.30(0.26) | 7.13(0.26) | 0.636 | t=0.37 |
| PHQ-9 | 6.50 (3.63) | 7.95 (3.76) | 10.72 (6.04) | 0.0050 | F=5.64 | 6.26(0.74) | 9.83(1.18) | 0.014 | t=2.57 | 10.52(0.92) | 10.70(1.44) | 0.919 | t=0.10 |
| GAD-7 | 4.64 (3.71) | 5.77 (3.26) | 9.00 (4.84) | <0.001 | F=8.21 | 4.52(0.81) | 7.83(1.05) | 0.017 | t=2.49 | 8.70(0.63) | 7.96(1.20) | 0.588 | t=0.55 |
| Post-intervention current sensing intensity | NA | NA | NA | NA | NA | NA | NA | NA | NA | 2.07(0.21) | 2.07(0.23) | >0.999 | t<0.001 |

IGD: Internet gaming disorder; RGU: recreational game users; HC: healthy controls; QGU-B: Questionnaire of Gaming Urges-Brief; IGDS9-SF: 9-item Internet gaming disorder scale-short form; DSM-5: Diagnostic and Statistical Manual of Mental Disorders, Fifth Edition; PHQ-9: Patient Health Questionnaire-9; GAD-7: Generalized Anxiety Disorder-7, NA: not applicable.

### EEG characterization of IGD

In D1, resting-state EEG analysis revealed significant group differences in the whole-brain delta (F_(2, 2139)_ = 22.42, p < 0.001), theta (F_(2, 2139)_ = 30.15, p < 0.001) and alpha (F_(2, 2139)_ = 20.78, p < 0.001) and band power spectral density (PSD) (appendix pp 4, Figure S1).

During gaming cue reactivity tasks, IGD participants exhibited significantly higher craving scores compared to participants from both HC (p < 0.001) and RGU groups (p = 0.003) under positive gaming cues. IGD patients also had higher craving scores for positive cues than for neutral cues (p < 0.001) or negative cues (p < 0.001) (Figure 2A). These results hence indicated that positive gaming cues specifically amplify craving in IGD.

**Figure 2:**
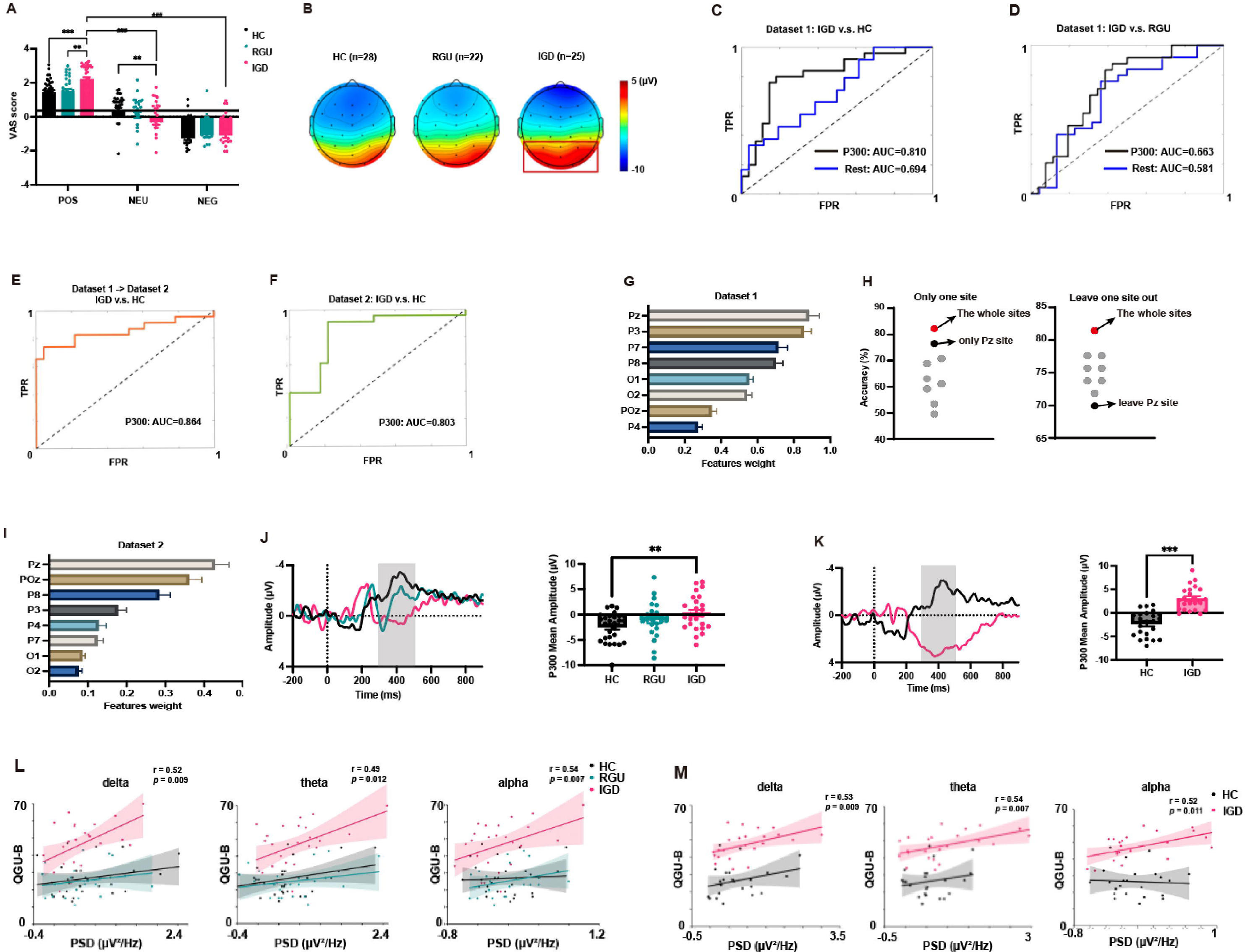
EEG-based machine learning model to identify IGD. A, B: The differences of the P300 components under game cues among three groups.A: The VAS craving score under different cues among 3 groups in game cue reactivity task. B: The topographic map of P300 component (300∼500ms) of parietal-occipital lobe under positive game cue among 3 groups. Red represents high amplitude, and blue represents low amplitude. EEG-based machine learning model to identify IGD and HC (C) or RGU (D) in Dataset 1. The blue line is the ROC curve of the model based on the energy of each frequency band at 31 electrode sites throughout the brain in the resting eye-closed state, the black line is the ROC curve of the machine learning model based on the energy of each frequency band of the 8 electrode points under the time window of the P300 of the parieto-occipital lobe under positive game cues. E, F: EEG-based machine learning model to identify IGD and HC in Dataset 2. E: Training on Dataset 1 and tested on Dataset 2. Weights of each electrode point in the machine learning model for identify IGD and HC based on the energy of each frequency band under positive game cue in Dataset 1 (G) and Dataset 2 (I).H: Accuracy by machine learning for identifying IGD and HC based on the energy of each frequency band under positive game cue with each electrode site separately and excluded in Dataset 1. The red points represented all the electrode sites, the black points represented Pz site, and the gray points represented other 7 electrode points. The P300 at Pz electrode site under positive game cue among three groups in Dataset 1 (J) and Dataset 2 (K). The horizontal axis is time (ms), the vertical axis is amplitude (μV), and the time point 0 ms is the time point at which the cues appeared. The shaded area is the time window (300-500 ms) in which the P300 component appeared. The associations between power in each frequency band and QGU-B craving scores at Pz in the P300 time window under positive game cue for three groups in Dataset 1 (L) and Dataset 2 (M). IGD: Internet gaming disorder; RGU: recreational game user; HC: healthy control; PSD: power spectral density; QGU-B: questionnaire of gaming urges-brief. ** p<0.01; *** p<0.001.

Parieto-occipital P300 component under positive cues differed significantly between IGD, RGU and HC (F_(2, 576)_ = 31.81, p < 0.001). Post-hoc analyses confirmed higher P300 amplitudes in IGD versus HC (p < 0.001) (Figure 2B). Analysis of P300 time-window PSD revealed group differences in theta (F_(2, 576)_ = 6.27, p = 0.002) and alpha (F_(2, 576)_ = 3.62, p = 0.028) bands (appendix pp 6, Figure S3).

Consistent with findings in D1, parieto-occipital P300 mean amplitudes were significantly higher in IGD versus HC (F_(1, 352)_ = 174.60, p < 0.001) in D2 (appendix pp 5, Figure S2A). PSD analysis identified group differences in delta (F_(1, 352)_ = 22.76, p < 0.001) and theta (F_(1, 352)_ = 37.91, p < 0.001) bands. (appendix pp 5, Figure S2B).

### Parieto-occipital P300 response to positive game cues optimally identified IGD and correlated with craving

SVM models using resting-state whole-brain spectral features achieved moderate accuracy (66.0%) in distinguishing IGD from HC in D1 (sensitivity: 70.8%, specificity: 61.5%, AUC: 0.718). In contrast, models incorporating time-window PSD features of parieto-occipital P300 under positive cues achieved superior accuracy (accuracy: 81.1%, sensitivity: 80.0%, specificity: 82.1%, AUC: 0.810) (Figure 2C). Similarly, classification of IGD versus RGU was more accurate using cue-induced P300 features than resting EEG features (AUC: 0.681 vs. 0.609) (Figure 2D).

Validation in D2 confirmed the robustness of the D1 P300-based model (accuracy: 80.9%, sensitivity: 82.2%, specificity: 78.3%, AUC = 0.864) for distinguishing IGD from HC (Figure 2E). Internal validation of the cue-induced P300-based model within D2 yielded comparable performance (accuracy: 78.3%, AUC = 0.801), demonstrating reproducibility (Figure 2F).

Weighted feature analysis identified the Pz electrode as the top contributor of the SVM model in both datasets (Figure 2G, I). Removing Pz features in D1 reduced IGD-HC classification accuracy by 11.3%, exceeding the impact of other electrodes (3.7∼9.4%), and using Pz alone achieved 76.5% classification accuracy, outperforming other single-electrode models (accuracy: 49.6∼69.8%) (Figure 2H). IGD had larger P300 component at Pz under positive cues (D1: F_(2, 72)_ = 4.67, p = 0.012; D2: t_(44)_ = 6.93, p < 0.001) (Figure 2J, K).

Spearman correlation analyses revealed strong positive associations between of Pz delta (D1: r = 0.52, p = 0.009; D2: r = 0.53, p = 0.009), theta (D1: r = 0.49, p = 0.012; D2: r = 0.54, p = 0.007), and alpha (D1: r = 0.42, p = 0.037; D2: r = 0.52, p = 0.011) power with QGU-B craving scores in IGD. No such correlations were observed in RGU or HC (p > 0.05) (Figure 2L, M).

### Participant characteristics of the intervention study

50 individuals with IGD were randomized into active (n=25) and sham (n=25) intervention groups, with 23 participants per group completing all interventions and follow-ups (total n=46) (Figure 1C). Baseline demographic and gaming-related measures—including age, sex, daily gaming time, QGU-B and VAS craving scores, IGDS9-SF, DSM-5 criteria, PHQ-9, and GAD-7 scores—showed no significant differences between active and sham groups (p > 0.05). Blinding efficacy was confirmed, as subjective current sensation ratings did not differ between tw<u>o</u> groups (p > 0.05) (Table 1). Both groups exhibited significant reductions in positive (time main effect: F_(2, 88)_ = 9.98, p < 0.001) and negative (time main effect: F_(2, 88)_ = 29.47, p < 0.001) mood over time (appendix pp 7, Figure S4). However, no significant group main effects or group × time interactions were observed for anxiety or depression scores (appendix pp 8, Figure S5), indicating that tDCS did not specifically modulate emotional states.

### Effects of intervention on the craving and gaming time of IGD

Compared with the sham group, active tDCS significantly reduced craving (QGU-B: F_(6, 264)_ = 13.26, p < 0.001; VAS: F_(6, 264)_ = 8.52, p < 0.001). Post-hoc analyses revealed lower craving in the active group at post_1 week (QGU-B: p = 0.043), post_2 week (QGU-B: p = 0.007; VAS: p = 0.046), and post_3 week (QGU-B: p = 0.010; VAS: p = 0.041) (Figure 3A, B). Daily gaming time decreased significantly in the active group versus sham (F_(4, 176)_ = 6.58, p < 0.01), with significant post-hoc reductions at post_3 week (p = 0.011) and post_4 week (p = 0.002) (Figure 3C).

**Figure 3:**
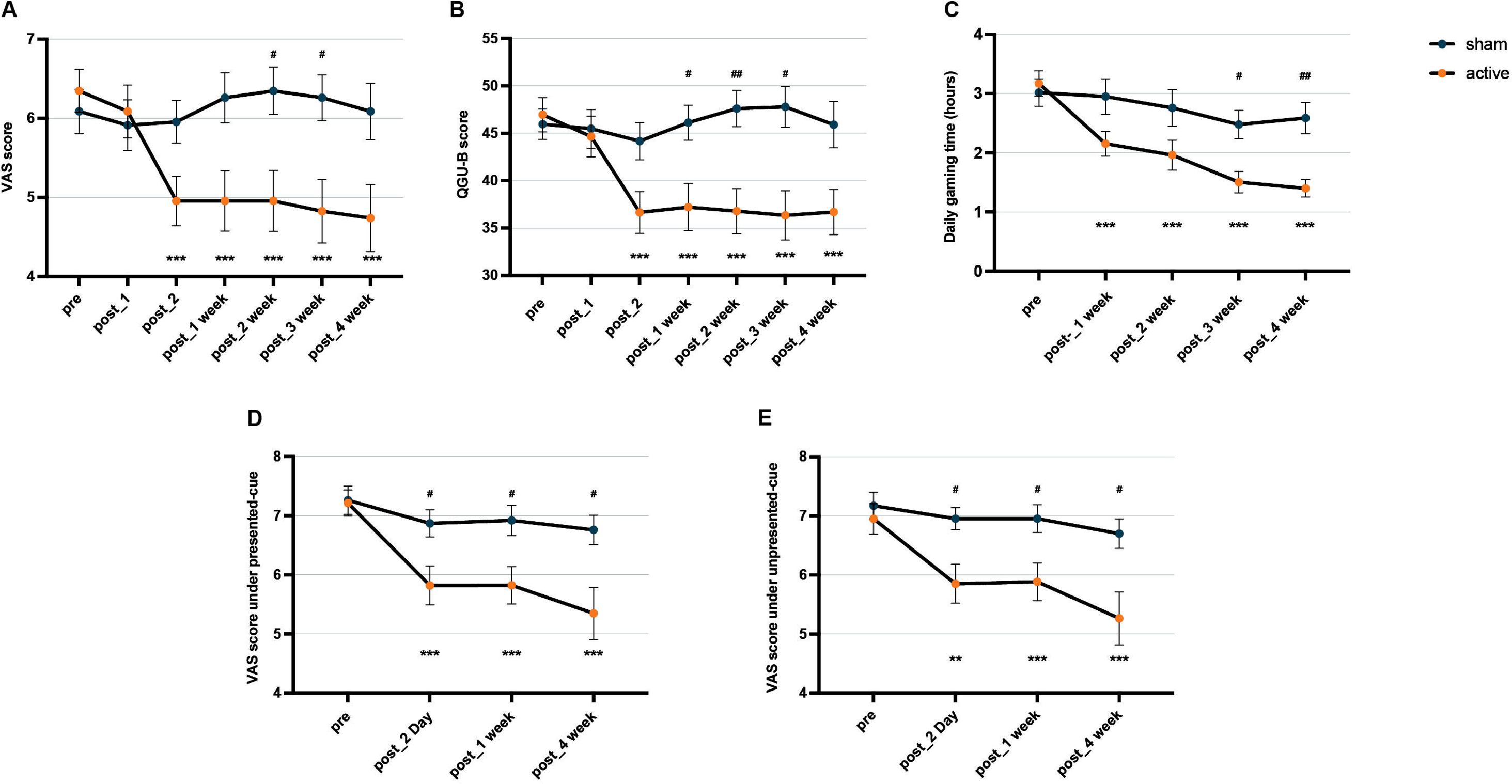
The effects of intervention on cravings and playtime in IGD. A: The changes of VAS score of craving. B: The changes of QGU-B score. C: The changes of daily gaming time. The changes of VAS score of craving under presented (D) and unpresented (E) game cue. Active: active tDCS stimulation group; sham: sham tDCS stimulation group; QGU-B: questionnaire of gaming urges-brief; unpresented-cue: the game cues not presented during tDCS; presented-cue: the game cues presented during tDCS. *: active group at each time point compared to pre-intervention in post-hoc analysis; #: comparison between the two groups at each time point in post-hoc analysis; ** p<0.01; *** p<0.001; # p<0.05; ## p<0.01.

Then we assessed whether the effects of active tDCS could be generalized to the craving under presented and unpresented gaming cues. IGD subjects reported higher VAS craving for gaming-related cues versus neutral cues (F_(2, 132)_ = 139.60, p < 0.001) with no difference between presented and unpresented gaming cues at pre-intervention (appendix pp 10, Figure S7). Post-intervention, active tDCS decreased craving for both presented (F_(3, 132)_ = 5.08, p = 0.02) and unpresented (F_(3, 132)_ = 3.86, p = 0.011) gaming cues at post_2 Day (p = 0.049), post_1 week (p = 0.039), and post_4 week (p = 0.034) compared to sham (Figure 3D-E and appendix pp 11, Figure S8).

### Effects of intervention on cue-evoked P300 of IGD

No significant group × time interactions were observed for P300 amplitudes at Pz during exposure to presented gaming cues (appendix pp 9, Figure S6A). However, active tDCS significantly reduced P300 amplitudes for unpresented gaming cues (group × time interaction: F_(3, 132)_ = 2.87, p = 0.039), with post-hoc decreases at post_2 Day (p = 0.047), post_1 week (p < 0.001), and post_4 week (p = 0.046) compared to baseline. At post_1 week, P300 amplitudes of the active group were significantly lower than sham (p = 0.022) (appendix pp 9, Figure S6B). Spearman correlation analysis revealed a significant positive association between P300 amplitude reduction and craving reduction in the active group (r = 0.56, p = 0.006), but not in the sham group (appendix pp 9, Figure S6C).

Time-frequency analysis demonstrated that alpha power at Pz for presented cues was significantly decreased in the active group (group × time interaction: F_(3, 132)_ = 2.71, p = 0.048), especially at post_1 week (p = 0.011) (appendix pp 9, Figure S6D-F). For unpresented cues, active tDCS reduced delta (group × time interaction: F_(3, 132)_ = 2.75, p = 0.045) and theta (group × time interaction: F_(3, 132)_ = 2.71, p = 0.048) power at post_1 week (delta: p = 0.006; theta: p = 0.029) and post_4 week (delta: p = 0.049) (appendix pp 9, Figure S6G-I and appendix pp 15, Figure S12).

Notably, baseline delta power at Pz during gaming cues emerged as the sole significant predictor of craving reduction at 4-week follow-up among P300 components, showing inverse correlations with both VAS (r = –0.58, p = 0.004) and QGU-B scores (r = –0.60, p = 0.003) (appendix pp 16, Figure S13). This suggests individuals with lower pre-intervention delta power achieved greater craving reduction.

### Effects of intervention on cognitive performance of IGD

No differences in cognitive performance (SST, MID, IGT) were observed between the active and sham tDCS groups at pre-intervention (appendix pp 3, Table S1). In SST, no significant group × time interactions were found for Go trial accuracy, Go reaction time, Stop trial accuracy, or Stop trial reaction time (appendix pp 12, Figure S9). In MID task, also no significant group × time interactions were found for sensitivity (d’) and reaction time in all conditions (appendix pp 13, Figure S10). Interestingly, in IGT, significant group × time interactions emerged for net score at post_1 week (F_(5, 220)_ = 2.39, p = 0.039) and post_4 week (F_(5, 220)_ = 2.85, p = 0.016). Post-hoc tests indicated that the active group exhibited a higher net score than the sham at post_1 week (Block 1: p = 0.013) and post_4 week (Block 6: p = 0.049), demonstrating progressive decision-making enhancement (appendix pp 14, Figure S11).

## Discussion

This study employed machine learning to identify EEG biomarkers for IGD and evaluated the effectiveness of targeted tDCS interventions based on these neural signatures. We established the parieto-occipital P300 component (IGD>HC) during positive game-cue exposure as a robust electrophysiological marker for both IGD diagnosis and craving. Our randomized, double-blind, sham-controlled trial revealed that Pz-targeted cathodal tDCS synchronized with simultaneously cue exposure significantly reduced craving, gaming time, cue-induced parietal hyperactivity and decision-making abnormality in IGD patients, with treatment effects persisting at one-month follow-up. These results provide a foundation for developing precision neuromodulation therapies for behavioral addictions.

Previous resting-state EEG studies on IGD have reported inconsistent results, including increased and decreased beta/theta power compared to healthy controls.^27,28^ Our machine learning models revealed that resting-state EEG features alone achieved only moderate classification accuracy (60%) for IGD diagnosis, significantly lower than cue-evoked EEG features (81.1% accuracy). Although resting-state beta power effectively identifies methamphetamine addiction and predicts craving, and distinguishes IGD from alcohol addiction, our results indicate its limited specificity for IGD, emphasizing the importance of task-based paradigms to capture disorder-specific neural responses. ^29,30^

The visual-evoked P300 component, generated from parietal cortex activity, reflects attentional allocation and working memory processes.^31^ Addiction researches consistently demonstrate enhanced P300 amplitudes under drug cues (e.g., cannabis, nicotine, and opioids), indicating attentional bias toward addiction-related stimuli. ^32–34^ Similarly, fMRI studies reveal increased parieto-occipital activation during gaming cues in problematic gamers.^35^ Our findings demonstrate that enhanced parietal P300 responses to gaming cue exposure serve as a reliable neurophysiological biomarker of IGD, marking dysregulated attentional processing of gaming-related stimuli.

P300 event-related oscillations (EROs) primarily comprise delta and theta bands. Studies have reported altered theta/delta activity in alcohol dependence during the P300 window.^36,37^ Alpha-band desynchronization within this window reflects cognitive resource engagement, as observed in food cues responses among overweight individuals.^38,39^ Our machine learning models confirmed that Pz delta, theta, and alpha oscillations as craving-corelated biomarkers across datasets. These findings align with prior reports linking P300 amplitudes to opioid craving and alpha oscillations to food craving.^32,39^ These findings parallel nicotine addiction studies where P300 normalization predicted craving reduction, positioning parietal EROs as both diagnostic markers and therapeutic targets.^40^ Crucially, baseline P300 delta-band power predicted treatment response, highlighting its potential as a prognostic biomarker for personalized IGD neuromodulation.

While DLPFC modulation remains a mainstay of tDCS therapy, our findings highlight the unique value of parietal cortex stimulation for IGD. As a hub integrating sensory, motor, cognitive processes and attention networks, parietal regions respond well to neuromodulation.^41,42^ Prior studies show that anodal tDCS over parietal regions improves attention and working memory in healthy individuals, clinical symptoms in schizophrenia, and attention deficits in attention-deficit/hyperactivity disorder.^43–45^ Our cathodal parietal tDCS significantly reduced gaming-cue attentional bias, suggesting normalized attention allocation in IGD. This attention-focused modulation approach may extend interventions for other substance and behavioral addictions. Notably, our study addressed a key limitation in prior addiction research by including around 50% female participants, significantly improving gender balance over prior IGD tDCS trials.^18,46^ This enhances the generalizability of our findings and provides valuable insights into potential gender-specific factors in IGD neuromodulation.

This study has several limitations. First, the machine learning models’ generalizability may be constrained by moderate sample sizes. Second, participants were predominantly adult university students playing Honor of Kings, limiting extension to adolescents and diverse gaming populations. Third, task-state EEG showed limited power in distinguishing IGD from recreational gamers, suggesting future studies should incorporate multimodal neuroimaging to separate reward, attention and default-mode networks. Fourth, absence of a DLPFC stimulation group precludes direct comparison of parietal versus frontal tDCS efficacy. Finally, therapeutic effects might be enhanced through prolonged or personalized tDCS protocols.

In summary, this study identified the cue-evoked parieto-occipital P300 component as a robust electrophysiological marker for IGD diagnosis and craving assessment. Our randomized control trial demonstrated that Pz-targeted cathodal tDCS synchronized with gaming cue exposure produced significant, sustained (≥1 month) reductions in gaming time, craving and pathological neural activation of IGD. These findings establish cue-synchronized tDCS as an effective intervention approach and position the parietal cortex as a novel therapeutic target for IGD neuromodulation.

## Panel: Research in context

### Evidence before this study

Craving is a pivotal component in the addiction process. Intervening against craving is an effective way to treat addictive behaviors. In Internet gaming disorder (IGD), game-cue-induced craving underlies excessive gaming. Current interventions for gaming craving mainly use cognitive-behavioral therapy, with few effective physical interventions. But physical interventions can precisely modulate craving-related neural mechanisms. We conducted a systematic search of PubMed, the Cochrane Library, PsycINFO and Embase for articles without language restrictions from database inception to July 6, 2025, with the terms “transcranial direct current stimulation”, “tDCS”, “gaming disorder”, “Internet gaming disorder” and “online gamer”. 7 papers were identified. Previous transcranial direct current stimulation (tDCS) studies on IGD mostly applied 2 mA current for 20-30 minutes (left dorsolateral prefrontal cortex (DLPFC) as anodal, right DLPFC as cathodal), with about 10 sessions. This approach consistently reduced gaming craving but had inconsistent effects on other cognitive abilities. Limitations included mostly male participants and small sample sizes (n<34). Our study involved 46 IGD patients of both genders. We firstly identified the cue-evoked parieto-occipital P300 as a definitive electrophysiological marker for IGD diagnosis (>80% accuracy) by using machine learning across two datasets. Then, based on this, we performed a randomized double-blind trial targeted tDCS intervention with the cathode at the Pz electrode (23 IGD for sham and 23 IGD for active, with 2mA current for 20 minutes within 2 consecutive days), showing promising efficacy.

### Added value of this study

First, machine learning analyses across two independent datasets identified and validated the parietal P300 elicited by gaming cues as the optimal EEG biomarker for IGD diagnosis and craving. This biomarker demonstrated higher discriminative accuracy than resting-state EEG features reported in prior literature.

Second, this study implemented novel cathodal tDCS targeting the parietal Pz site during gaming cue exposure. The intervention significantly reduced craving (p<0.001) and gaming time (p<0.001) immediately post-treatment and throughout the 1– to 4-week follow-up period in the active group. These effects were accompanied by improved reward sensitivity and decision-making, establishing the parietal cortex as a promising neuromodulation target alternative to the DLPFC for IGD treatment.

### Implications of all the available evidence

These findings advance precision IGD biomarkers and evidence-based neuromodulation. They provide validated neurophysiological biomarkers for IGD identification and demonstrate, via randomized controlled trial, efficacy of novel EEG-targeted tDCS. This establishes cue-synchronized tDCS as an effective therapy and positions the parietal cortex as a novel neuromodulation target – offering innovative approaches for IGD prevention and intervention.

## Contributors

Y.S. and Y-X.S. conceived and designed the study, contributed to data analysis and interpretation, and wrote and revised the manuscript. Y-X.S., Y-C.H., Q-H.Z. contributed to data acquisition, processing, analysis and interpretation, and wrote and revised the manuscript. X-Q.Y., H-X.G. contributed to data acquisition and processing, and revised the manuscript. Y.G. contributed to data acquisition and revised the manuscript. J-J.B., G-D.Z., T-Y.J., J.S. contributed to data analysis and interpretation, and revised the manuscript. All authors contributed to the article and approved the final version of the manuscript.

## Data sharing

Data can be requested from the corresponding author. Data use and requests underlie the publication policy of the multicentre GET. FEEDBACK.GP-RCT.

## Acknowledgments

This work was supported by the STI2030-Major Projects (2022ZD0211200).

