## Supplementary Materials for "Effects of Parietal Cathodal tDCS during Game Cue Exposure on Internet Gaming Disorder: A Randomized Double-Blind Sham-Controlled Trial"

**Supplementary material**

**Contents**

Cognition paradigm testing	**2**

Table S1: Pre-intervention Cognitive indicators in two groups	3

Figure S1: The power of each frequency band of three groups at resting state in D1	4

Figure S2: The P300 EEG component under game cues in 2 groups in D2	5

Figure S3: The topographic map of P300 component (300~500ms) band power under positive game cue among 3 groups.	6

Figure S4: The change of positive and negative mood after tDCS in 2 groups	7

Figure S5: The change of depression and anxiety after tDCS in 2 groups	8

Figure S6: The effects of intervention on cue-evoked P300 in IGD	9

Figure S7: The pre-intervention VAS score of craving under different cues in IGD subjects of the intervention study	10

Figure S8: The effects of intervention on the VAS score of craving under game cues across two intervention days	11

Figure S9: The effects of intervention on cognitive performances in SST	12

Figure: S10 The effects of intervention on cognitive performances in MID	13

Figure S11: The effects of intervention on cognitive performances in IGT	14

Figure S12: The effects of intervention on cue-evoked P300 band power in IGD	15

Figure S13: Association of delta band power under game cues pre-intervention with intervention-decreased craving	16

References	17

**Cognition paradigm testing**

(1) The game cue reactivity task (GCRT) primarily tests the subjects’ craving level for game-related cues. In the task, a series of pictures is displayed on the screen, and each picture is scored on a VAS for game craving. The research exploration part includes 30 positive game cue pictures, 30 negative game cue pictures, and 30 neutral pictures; the intervention part includes 50 positive game cue pictures and 30 neutral pictures. Only positive game cue pictures are presented during the tDCS intervention, with a total of 30 presented; these are referred to as the game cue pictures presented in the intervention.

(2) The stop signal task (SST) is a classical experimental paradigm that assesses attentional and inhibitory control by measuring an individual’s ability to inhibit an initiated action during a task.^1^ Subjects are presented with two types of arrows facing left and right during the task, and are asked to press the key as soon as possible when the arrows appeared. After the appearance of some left and right arrows, an up arrow appears, requiring to stop the ongoing reaction immediately. The left and right arrows in the task are the “Go signals”, and the up-arrow appears immediately after the left and right arrows as the “Stop signals”. Behavioral indicators focus on the correct response rate of Go and Stop signals, the average response time of correctly responded Go signals, and the Stop Signal Reaction Time (SSRT).

(3) The monetary incentive delay task (MID) is a widely used experimental paradigm for studying subjects' behavioral performance in anticipation of and in response to reward and punishment, and the task is widely used in addiction research.^2^ The MID task involves three sequential phases: during the cueing phase, subjects view a cue indicating whether the upcoming trial offers a potential reward (monetary gain: large/¥5 or small/¥0.2 for success), punishment (monetary loss: large/¥5 or small/¥0.2 for failure), or neutral (no outcome). Subsequently, in the target phase, after a variable delay, a target stimulus appears requiring a rapid button press, with its duration dynamically adjusted to maintain approximately 50% success rate. Finally, the feedback phase displays the outcome (reward gained/avoided or punishment incurred/avoided) based on reaction speed. Key behavioral metrics include reaction time (from target onset to response) and accuracy (proportion of successful responses), with accuracy sensitivity further calculated as d' = Z (target stimulus accuracy) – Z (neutral stimulus accuracy), where Z represents the inverse of the standard normal cumulative distribution function.

(4) The Iowa Gambling Task (IGT) was employed to assess decision-making due to its real-world relevance. ^3,4^ In the task, subjects start with ¥200 and make 60 card selections across six stages (10 choices each) from four decks (A/B/C/D) to maximize gains. Decks A and B are disadvantageous (“loss decks”), offering high immediate rewards (¥10) but larger penalties (¥-25), while decks C and D are advantageous (“profit decks”) with lower rewards (¥5) but smaller penalties (¥-5). Through trial-by-trial feedback, participants learn to identify optimal decks. The primary behavioral metric is the net score per stage, calculated as (C + D choices) minus (A + B choices), where higher scores reflect better decision-making by favoring long-term advantageous options.

**Table S1:** **Pre-intervention Cognitive indicators in two intervention groups.**

|  | **Cognitive indicators** | **Active (n=23)** | **Sham (n=23)** | ***p*** | **Statistics** |
| --- | --- | --- | --- | --- | --- |
| SST | Go accuracy | 0.98 (0.004) | 0.97(0.01) | 0.348 | t=0.95 |
|  | Go reaction time (ms) | 412.52 (17.77) | 376.38 (12.14) | 0.107 | t=1.65 |
|  | Stop accuracy | 0.60 (0.02) | 0.63(0.02) | 0.398 | t=0.85 |
|  | Stop reaction time (ms) | 336.95 (8.46) | 317.32 (6.11) | 0.069 | t=1.87 |
| MID | Large win reaction time (ms) | 215.49(5.67) | 206.48(4.60) | 0.224 | t=1.23 |
|  | Little win reaction time (ms) | 213.63(4.88) | 204.96(4.48) | 0.198 | t=1.31 |
|  | Large loss reaction time (ms) | 214.07(5.03) | 204.98(4.63) | 0.191 | t=1.33 |
|  | Little loss reaction time (ms) | 213.17(3.06) | 208.65(3.49) | 0.335 | t=0.97 |
|  | The d’ of large win | 0.46(0.08) | 0.65(0.09) | 0.117 | t=1.60 |
|  | The d’ of little win | 0.35(0.08) | 0.36(0.09) | 0.877 | t=0.16 |
|  | The d’ of large loss | 0.36(0.10) | 0.49(0.10) | 0.384 | t=0.88 |
|  | The d’ of little loss | 0.26(0.09) | 0.30(0.07) | 0.662 | t=0.44 |
| IGT | Block 1 net scores | -2.32(0.61) | -1.83(0.63) | 0.621 | F=0.70 |
|  | Block 2 net scores | 0.23 (0.78) | -0.57 (0.86) |  |  |
|  | Block 3 net scores | -1.55(1.01) | -0.83(0.88) |  |  |
|  | Block 4 net scores | -0.14(0.84) | -0.83(0.83) |  |  |
|  | Block 5 net scores | 0.18(0.91) | -0.74(0.74) |  |  |
|  | Block 6 net scores | -0.55(0.99) | 0.17(1.01) |  |  |

SST: Stop signal task; MID: Monetary incentive delay task; IGT: Iowa gambling task; active: active tDCS stimulation group; sham: sham tDCS stimulation group; d’: D prime.

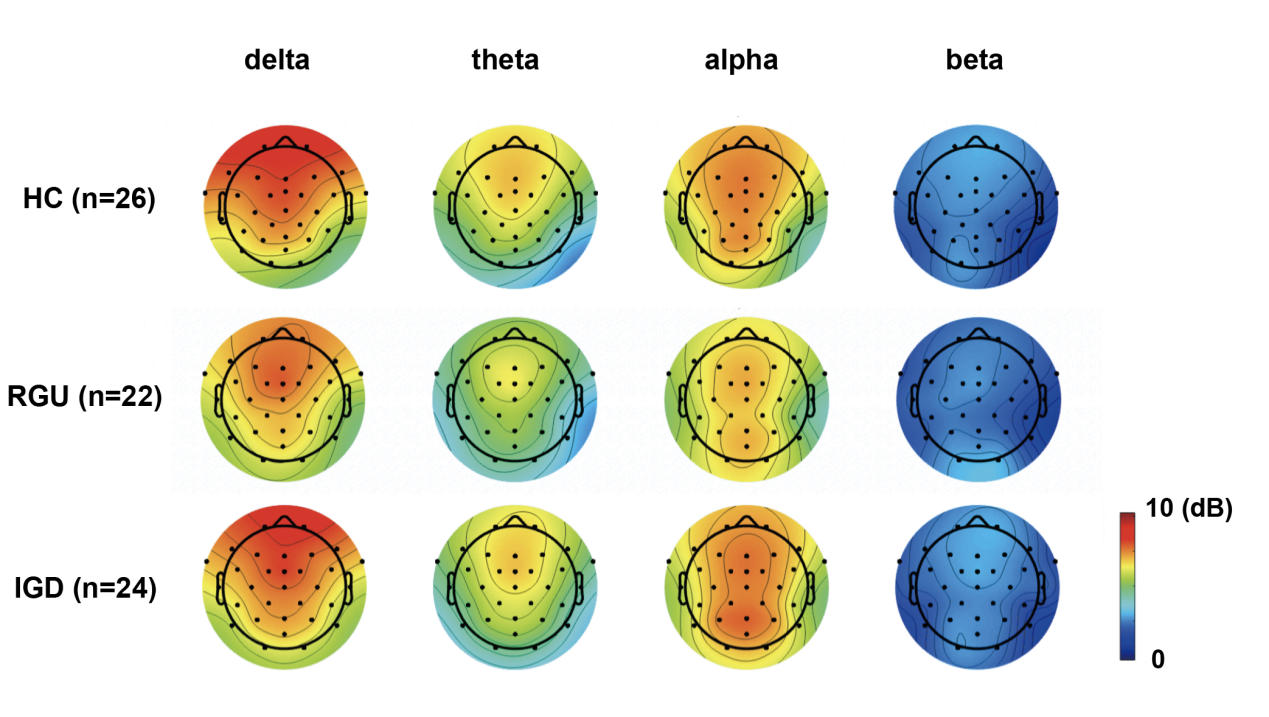
**Figure S1:** **The power of each frequency band of three groups at resting state in Dataset 1 of the exploratory study.**

Red indicated high power and blue indicated low power. IGD: Internet gaming disorder; RGU: recreational game users; HC: healthy controls.

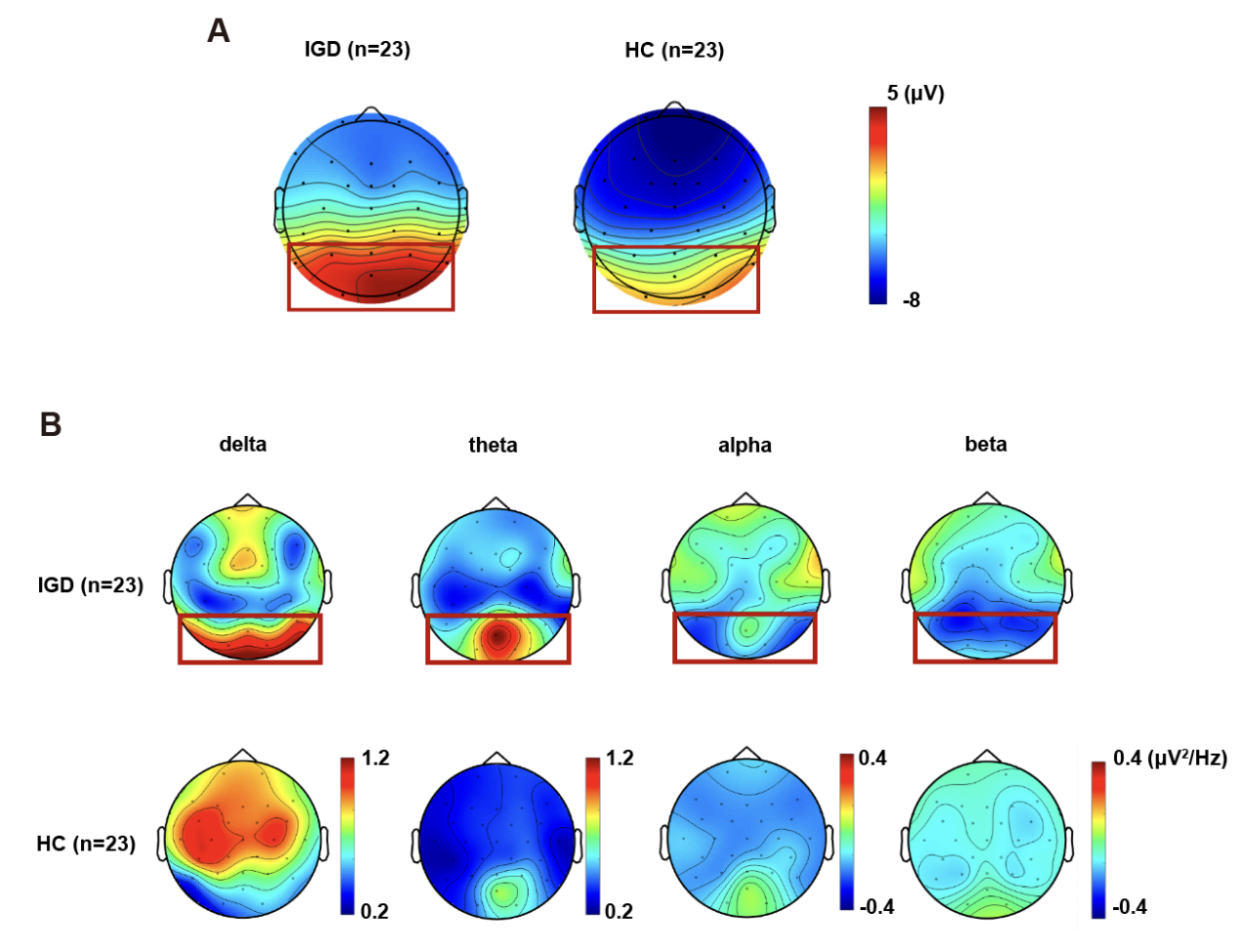

**Figure S2: The P300 EEG component under game cues in 2 groups in Dataset 2 of the exploratory study.**

A The topographic map of P300 component (300~500ms) of parietal-occipital lobe under positive game cue among 3 groups. Red represents high amplitude, and blue represents low amplitude. B The topographic map of P300 component (300~500ms) band power under positive game cue among 2 groups. Red represents high power and blue represents low power. The red boxes showed the electrode sites with P300 components in IGD group, which includes a total of 8 electrode sites in the parieto-occipital lobe portion (P3, P4, P7, P8, O1, O2, Pz, POz). IGD: Internet gaming disorder; HC: healthy controls.

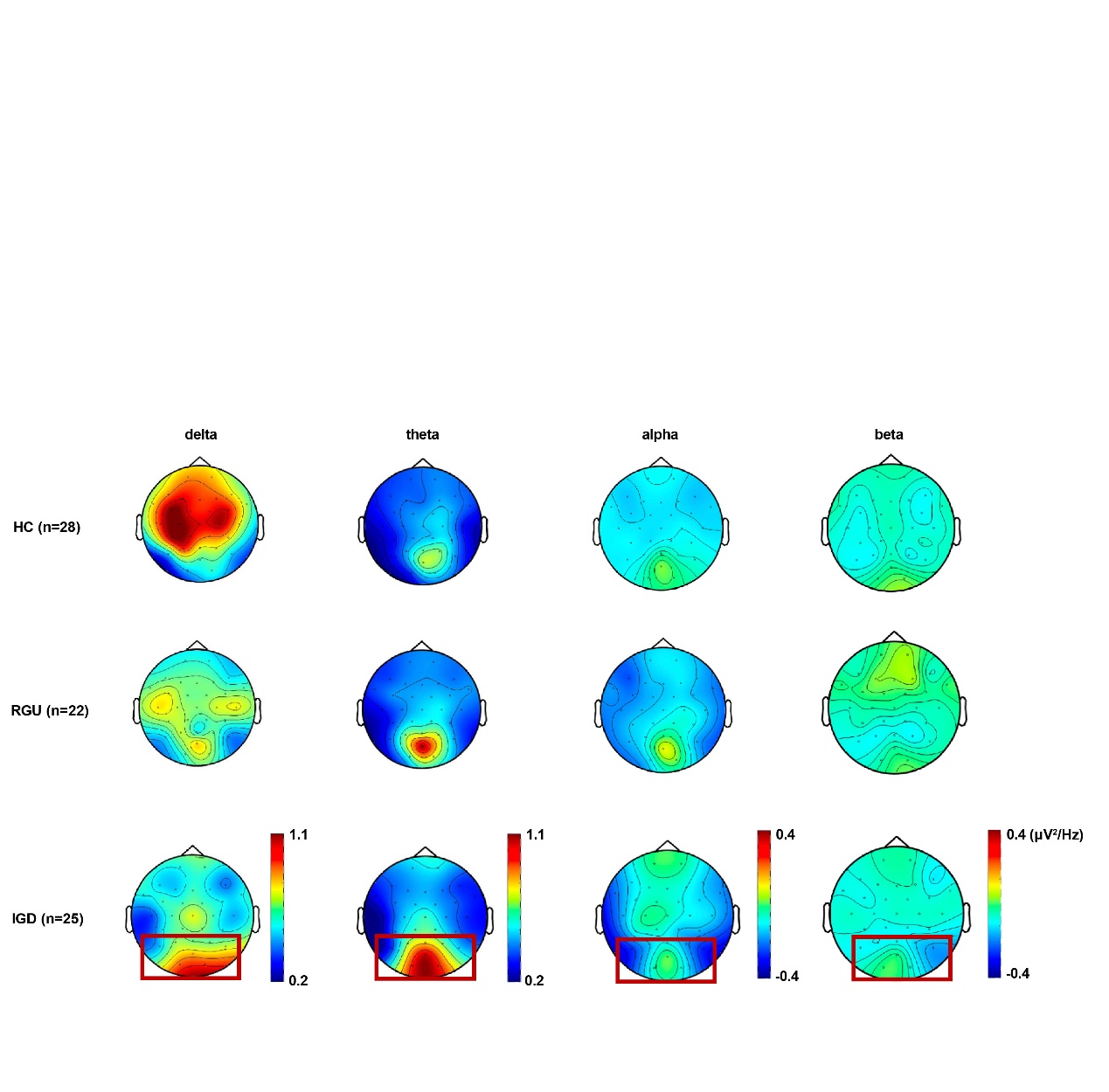
**Figure S3: The topographic map of P300 component (300~500ms) band power under positive game cue among 3 groups.**

Red represents high energy and blue represents low energy. The red boxes showed the electrode sites with P300 components in IGD group, which includes a total of 8 electrode sites in the parieto-occipital lobe portion (P3, P4, P7, P8, O1, O2, Pz, POz). IGD: Internet gaming disorder; RGU: recreational game user; HC: healthy control. POS: positive gaming cues; NEG: negative gaming cues; NEU: neutral cues. *: Comparison of VAS craving ratings in the IGD group vs. the other two groups under each condition; #: Comparison of craving ratings for positive game cues vs. the other two cue types within the IGD group; **: p<0.01; ***: p<0.001; ###: p<0.001.

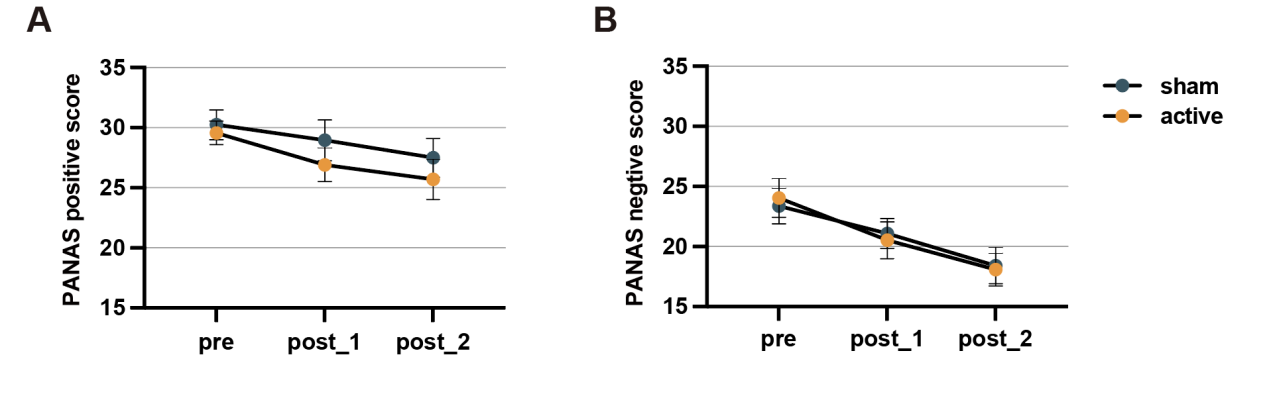

**Figure S4: The effects of intervention on positive and negative mood.**

Active: active tDCS stimulation group; sham: sham tDCS stimulation group; PANAS: Positive and Negative Affect Schedule.

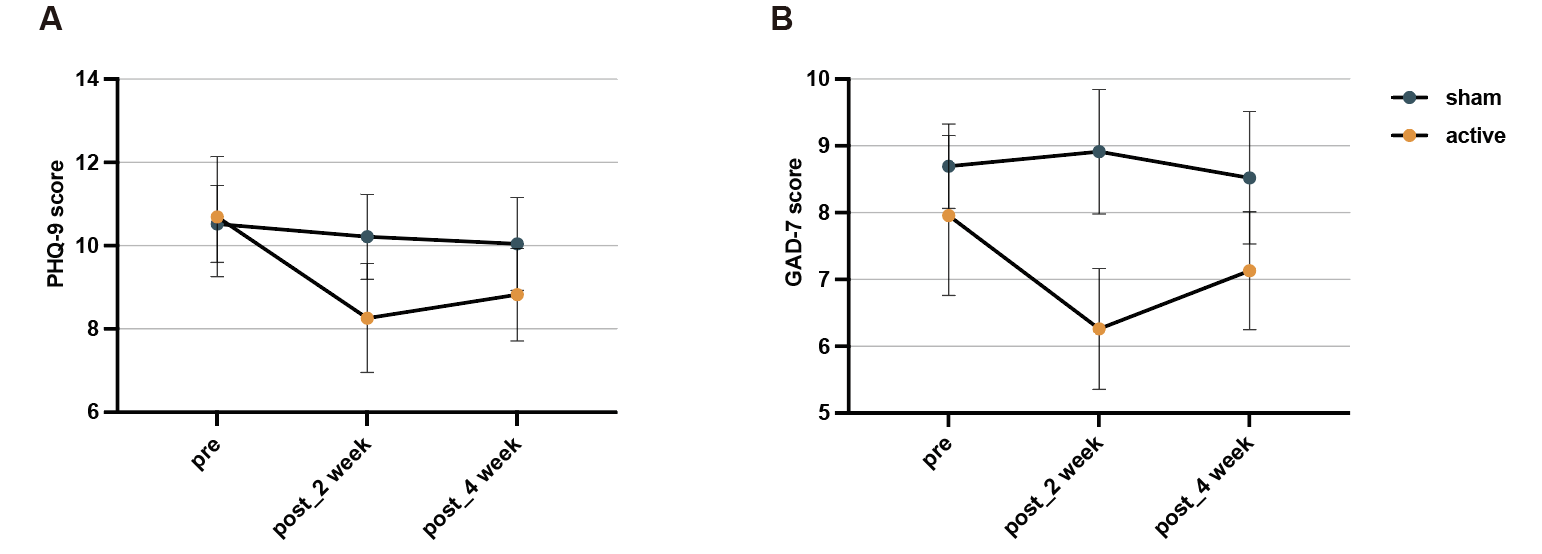

**Figure S5:** **The effects of intervention on depression and anxiety.**

Active: active tDCS stimulation group; sham: sham tDCS stimulation group; PHQ-9: Patient Health Questionnaire-9; GAD-7: Generalized Anxiety Disorder-7.

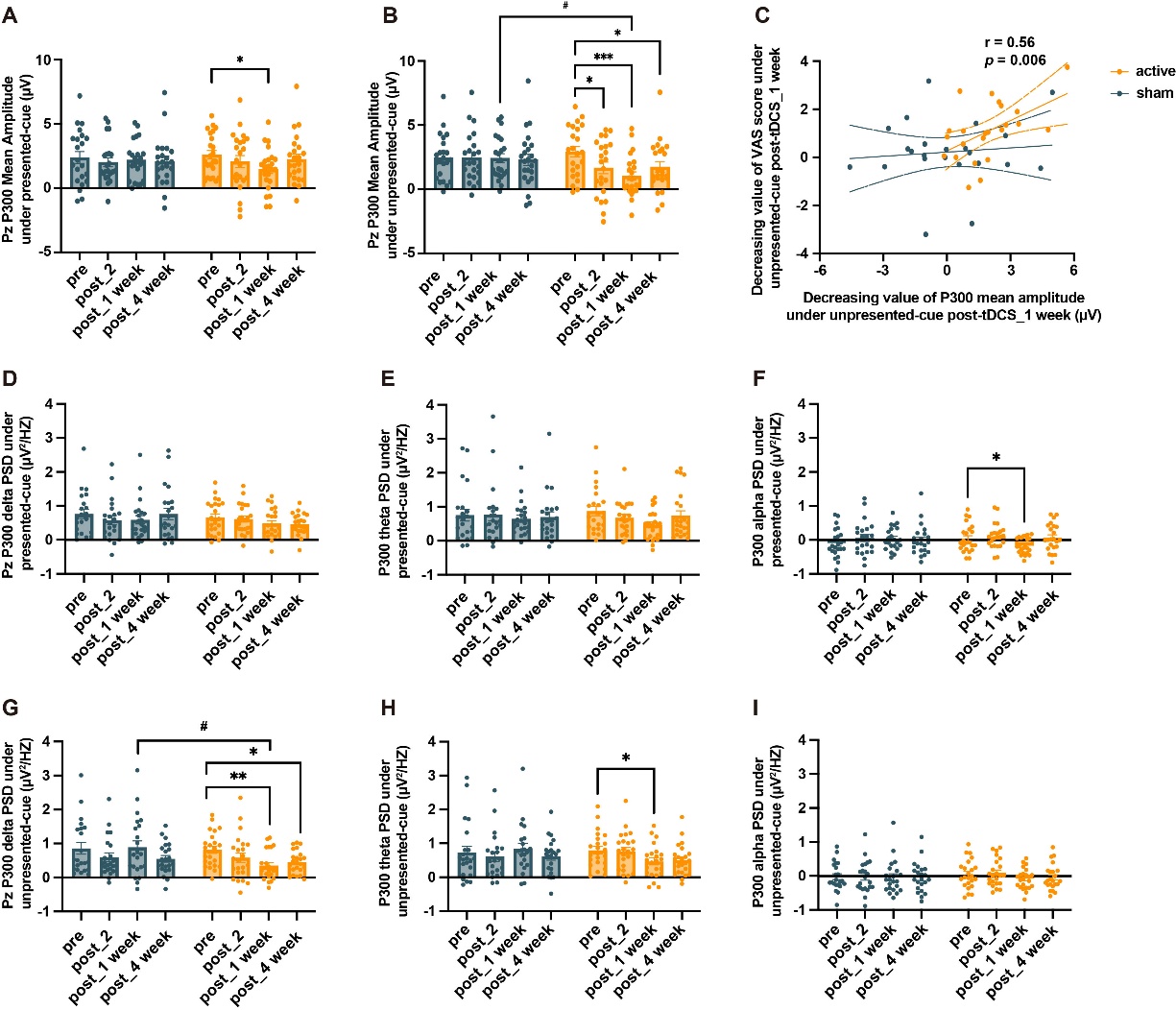
**Figure S6: The effects of intervention on cue-evoked P300 in IGD.**

The change of P300 mean amplitude under presented (A) and unpresented (B) game cues. C: The association between decreased P300 amplitude at Pz and decreased VAS craving score under unpresented game cues one week after tDCS. The change of P300 band power under presented (D~F) and unpresented (G~I) game cues. Active: active tDCS stimulation group; sham: sham tDCS stimulation group; unpresented-cue: the game cues not presented during tDCS; presented-cue: the game cues presented during tDCS. *: active group at each time point compared to pre-intervention in post-hoc analysis; #: comparison between the two groups at each time point in post-hoc analysis; * p<0.05; ** p<0.01; *** p<0.001; # p<0.05.

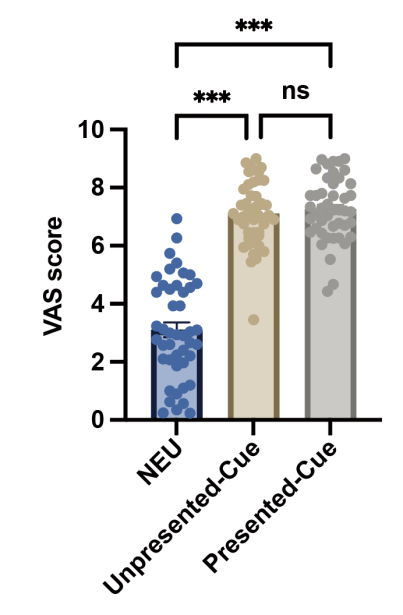

**Figure S7: The pre-intervention VAS score of craving under different cues in IGD subjects of the intervention study.**

NEU: neutral cues; Unpresented-Cue: the game cues not presented during tDCS; Presented-Cue: the game cues presented during tDCS. ***: p<0.001; ns: p>0.05.

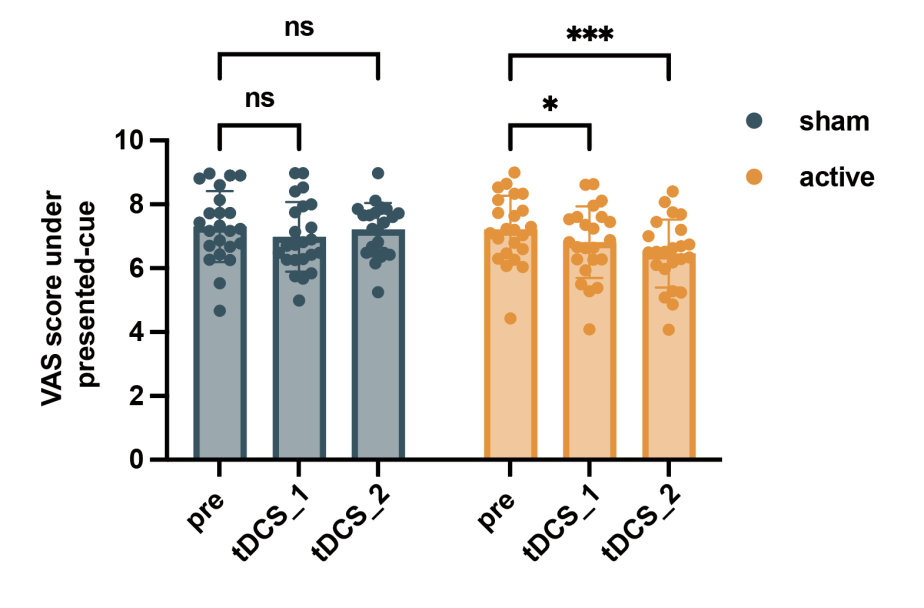

**Figure S8: The effects of intervention on the VAS score of craving under game cues across two intervention days.**

Active: active tDCS stimulation group; sham: sham tDCS stimulation group; presented-cue: the game cues presented during tDCS; tDCS_1: During the first day of the tDCS intervention; tDCS_2: During the second day of the tDCS intervention *p<0.05; ***: p<0.001; ns: p>0.05.

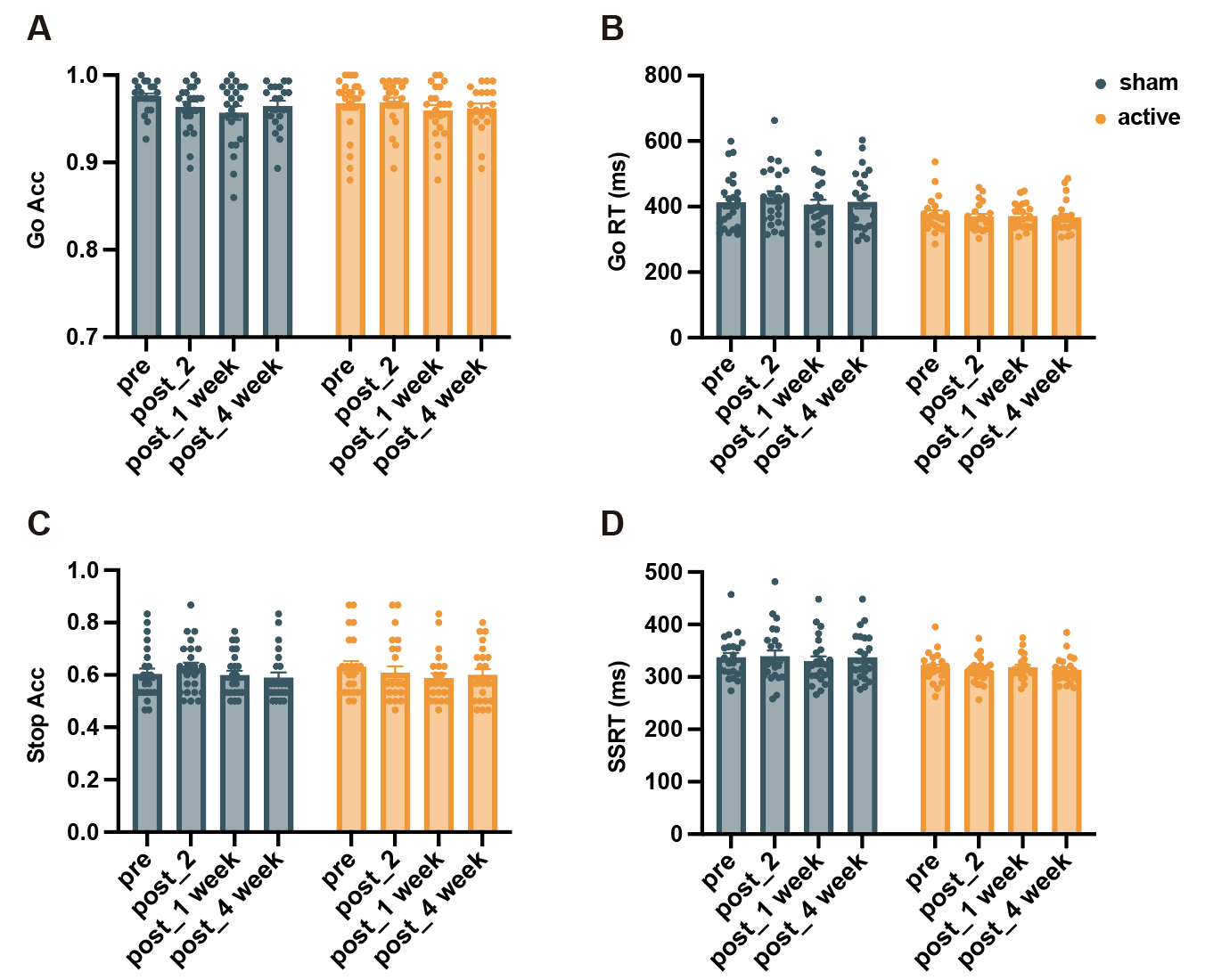
 **Figure S9: The effects of intervention on cognitive performances in SST.**

Active: active tDCS stimulation group; sham: sham tDCS stimulation group; SST: Stop signal task; Go Acc: Go accuracy; Go RT: Go reaction time (ms); Stop Acc: Stop accuracy; Stop RT: Stop reaction time (ms)

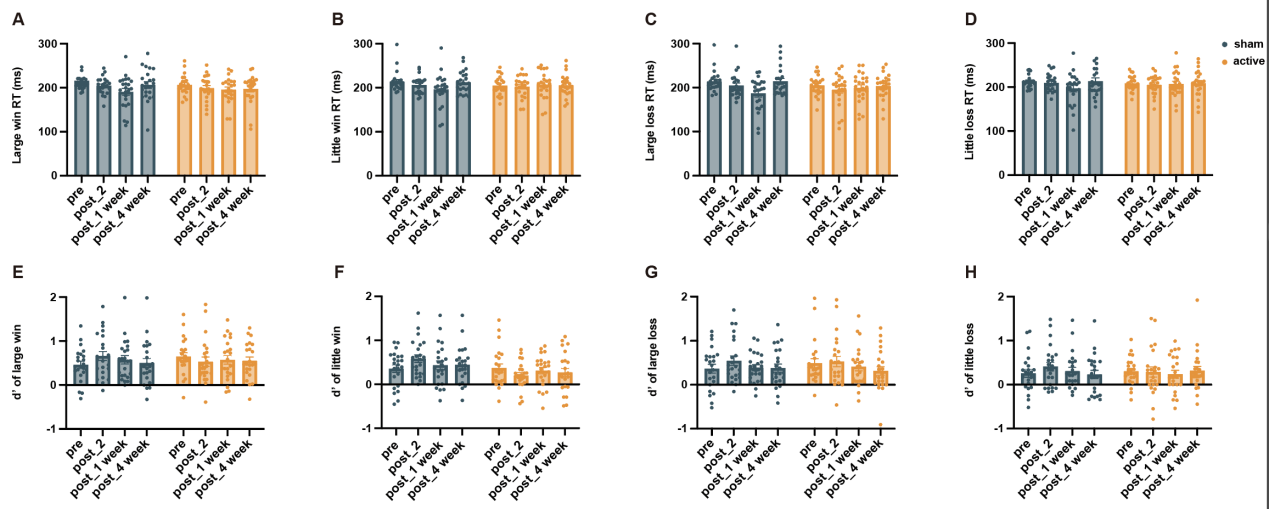

**Figure S10: The effects of intervention on cognitive performances in MID.**

Active: active tDCS stimulation group; sham: sham tDCS stimulation group; MID: Monetary incentive delay task; RT: reaction time (ms); d’: D prime.

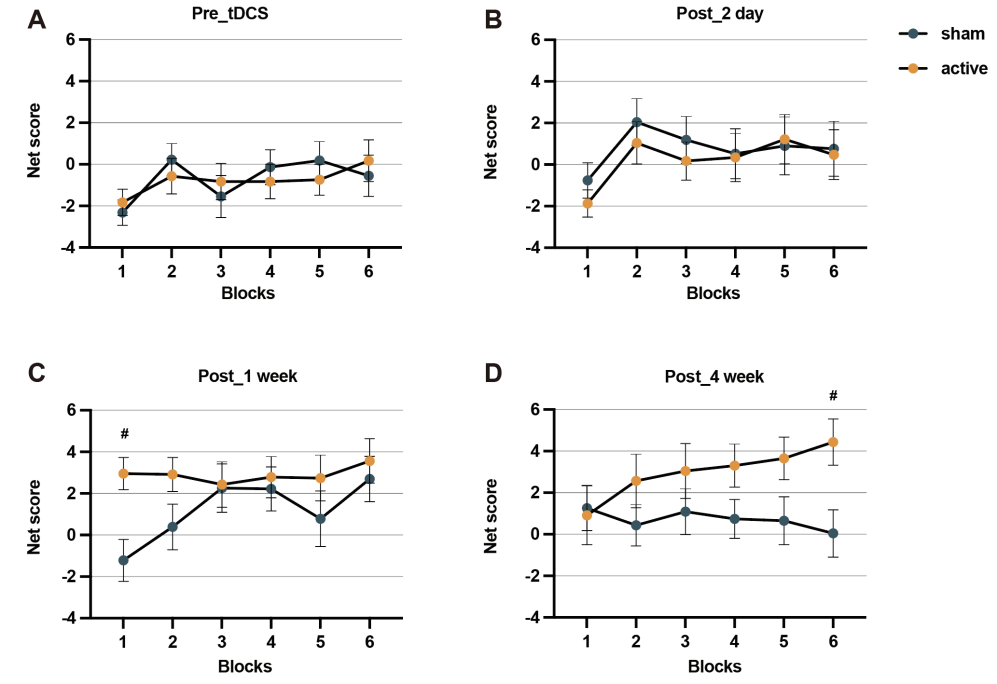

**Figure S11: The effects of intervention on cognitive performances in IGT.**

Active: active tDCS stimulation group; sham: sham tDCS stimulation group; IGT: Iowa gambling task. #: comparison between the two groups at each block in post-hoc analysis; #p<0.05.

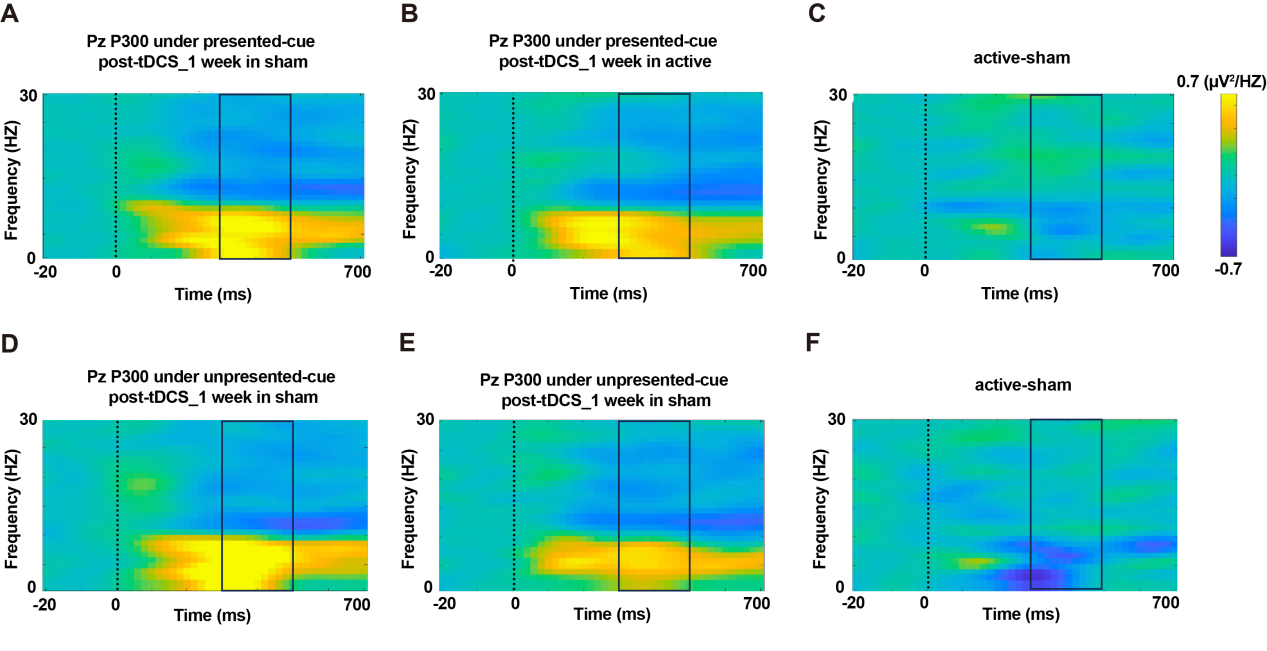

**Figure S12: The effects of intervention on cue-evoked P300 band power in IGD.**

Time-frequency energy maps at Pz under presented (A~C) and unpresented (D~F) game cues one week after tDCS for two groups, yellow represents high energy and blue low energy. The horizontal axis is time (ms), the vertical axis is frequency (Hz), and time point 0 ms is the time point at the cues appeared. The black boxes showed the time window when the P300 component appeared (300~500 ms). Active: active tDCS stimulation group; sham: sham tDCS stimulation group; unpresented-cue: the game cues not presented during tDCS; presented-cue: the game cues presented during tDCS.

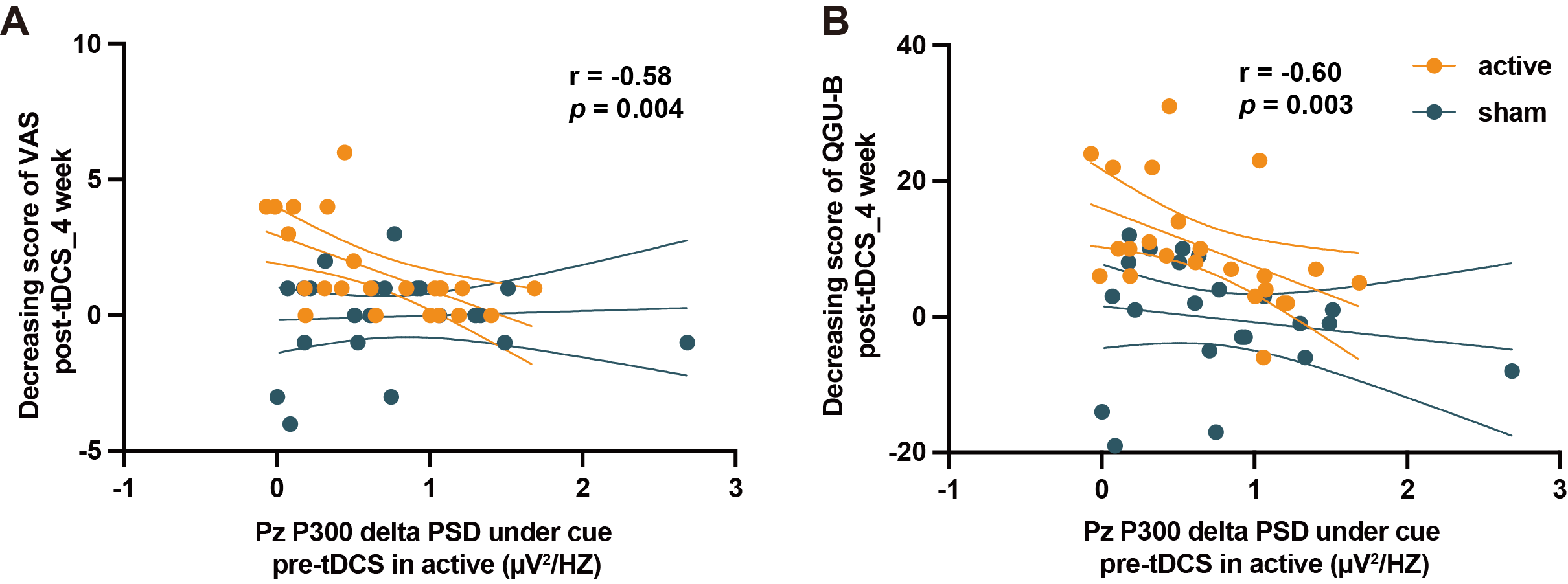

**Figure S13: Association of delta band power under game cues pre-intervention with intervention-decreased craving.**

The association of delta band power under game cues pre-intervention with craving change (A: VAS; B: QGU-B) four weeks post-intervention in two groups (active and sham groups). VAS: visual analogue scale; QGU-B: questionnaire of gaming urges-brief.
